# Early prediction of liver disease using conventional risk factors and gut microbiome-augmented gradient boosting

**DOI:** 10.1101/2020.06.24.20138933

**Authors:** Yang Liu, Guillaume Meric, Aki S. Havulinna, Shu Mei Teo, Matti Ruuskanen, Jon Sanders, Qiyun Zhu, Anupriya Tripathi, Karin Verspoor, Susan Cheng, Mo Jain, Pekka Jousilahti, Yoshiki Vazquez-Baeza, Rohit Loomba, Leo Lahti, Teemu Niiranen, Veikko Salomaa, Rob Knight, Michael Inouye

## Abstract

Gut microbiome sequencing has shown promise as a predictive biomarker for a wide range of diseases, including classification of liver disease and severity grading. However, the potential of gut microbiota for prospective risk prediction of liver disease has not been assessed. Here, we utilise shallow gut metagenomic sequencing data of a large population-based cohort (N=>7,115) and ∼15 years of electronic health register follow-up together with machine-learning to investigate the predictive capacity of gut microbial predictors, individually and in conjunction with conventional risk factors, for incident liver disease and alcoholic liver disease. Separately, conventional and microbiome risk factors showed comparable predictive capacity for incident liver disease. However, microbiome augmentation of conventional risk factor models using gradient boosted classifiers significantly improved performance, with average AUROCs of 0.834 for incident liver disease and 0.956 for alcoholic liver disease (AUPRCs of 0.185 and 0.304, respectively). Disease-free survival analysis showed significantly improved stratification using microbiome-augmented risk models as compared to conventional risk factors alone. Investigation of predictive microbial signatures revealed a wide range of bacterial taxa, including those previously associated with hepatic function and disease. This study supports the potential clinical validity of gut metagenomic sequencing to complement conventional risk factors for risk prediction of liver diseases.

## Introduction

Liver disease causes ∼2 million deaths per year worldwide, approximately 3.5% of all deaths, and is increasingly common in aging populations^[1, 2]^. The aetiology of liver disease is complex and includes several inter-related risk factors, such as obesity, age and excess alcohol consumption^[3]^. Alcohol consumption, in particular, is a major contributor to liver disease, accounting for >50% of cirrhosis deaths^[2]^. The consequences of liver disease can be acute or chronic with highly variable progression rates; however, most patients are not diagnosed until an advanced stage when liver function is overwhelmed (e.g. decompensated cirrhosis)^[4, 5]^. Currently, liver biopsy remains the gold standard for diagnosis and classification of disease stage, but biopsy is invasive and thus restricted. Although non-invasive tests for detecting liver disease are available, such as ultrasound, computed tomography, magnetic resonance imaging and spectroscopy, they are primarily applicable to the detection of advanced severity^[6-8]^. Hence, there is an unmet need for high fidelity early detection and risk prediction approaches for liver disease.

The role of the human gut microbiome—the collection of microorganisms residing in the gastrointestinal tract—has been increasingly recognized in various aspects of liver disease^[9, 10]^. Interest in the gut microbiome has rapidly grown as sequencing technologies have progressed from 16S rRNA amplicon sequencing to shotgun metagenomics. Recent studies have revealed evidence linking gut microbial composition and the pathogenesis of liver disease^[11-13]^, as well as potential therapeutic approaches targeting gut microbial communities^[14, 15]^. Importantly, the gut microbiome has shown potential for the differentiating cirrhosis and non-cirrhosis controls. Qin et al. showed gene and function level biomarkers derived from metagenomics could classify liver cirrhosis patients and healthy controls^[16]^. Loomba et al. successfully distinguished advanced fibrosis from mild and moderate NAFLD using gut microbiome characterized by whole-genome shotgun sequencing with random forest classifiers^[17]^. Later, Caussy et al. used random forest classifiers to distinguish NAFLD-cirrhosis from non-NAFLD healthy controls based on gut microbial compositions from 16S sequencing^[18]^. However, previous studies have been limited by cross-sectional study design and there are limited data regarding the longitudinal association between baseline microbiota and incident liver disease. This would be an important step in investigating whether the gut-microbiome is causally linked to liver disease or can be used as a stratification tool to identify those at high risk, who may benefit from targeted interventions.

Therefore, we designed a longitudinal study to examine the association and predictive capacity of the gut microbiome and incident liver diseases, using shallow metagenomic sequencing and supervised machine learning in a large population-based cohort of >7000 individuals with over 15 years of electronic health records (EHR) follow-up. Traditional statistical and machine learning approaches are compared on gut metagenomes, and their predictive capacity is evaluated individually and in combination with conventional risk factors, including age, sex, body mass index, waist-hip ratio, alcohol consumption, smoking status, triglycerides, high-density lipoprotein cholesterol, low-density lipoprotein cholesterol, and gamma-glutamyl transferase levels. The best performing models are further assessed using survival analysis for time to disease onset. Taken together, our study assesses the potential clinical validity for adding the gut metagenome to conventional risk factors for prediction of incident liver disease. We make our predictive models freely available (see **Data Availability**).

## RESULTS

### CHARACTERISTICS OF STUDY POPULATION

This study included 7115 participants with a median follow-up of 14.8 years from the population-based FINRISK 2002 cohort whose participants are representative of Finnish population aged 25-74 years at baseline (**Methods**)^[19]^. The detailed baseline characteristics of the study population are provided in

**Table 1**. To investigate the predictive capacity of baseline gut microbiome and conventional risk factors for incident liver diseases, we matched phenotype metadata with gut microbial profiles derived from stool samples, and linked the baseline data to follow-up diagnoses of any liver diseases (LD) or alcoholic liver disease (ALD) defined by ICD-10 codes (**Methods**). After stringent quality control and filtering (**Methods**), 41 cases of incident ALD and 103 cases of incident LD were considered for prediction analyses.

**Table 1.** Baseline characteristics of study population.

| N=7115 | Female | Male |
| --- | --- | --- |
|  | n=55% | n=45% |
| **Demographics** |  |  |
| Age | 49.69 [38.05, 58.78] | 51.92 [40.54, 60.70] |
| **Physical parameters** |  |  |
| Body mass index (kg/m<sup>2</sup>) | 25.90 [23.09, 29.47] | 26.9 [24.55, 29.58] |
| Waist-hip ratio | 0.84 [0.80, 0.88] | 0.97 [0.92, 1.01] |
| **Lifestyles** |  |  |
| Smoking | 19% | 28% |
| Pure alcohol consumption (g/week) | 18.9 [2.7, 55.8] | 75.9 [20.7, 168.3] |
| **Laboratory results** |  |  |
| HDL cholesterol (mmol/l) | 1.59 [1.35, 1.89] | 1.30 [1.10, 1.53] |
| LDL cholesterol (mmol/l) | 3.19 [2.65, 3.76] | 3.46 [2.89, 4.09] |
| Triglycerides (mmol/l) | 1.07 [0.80, 1.45] | 1.36 [0.97, 1.97] |
| Gamma-glutamyl transferase (U/L) | 19 [15, 27] | 30 [21, 46] |

Median [IQR] for continuous variables; n% for categorical variables

### BASELINE GUT MICROBIAL COMPOSITION

Stool samples were sequenced by shallow shotgun metagenomics to a mean depth of approximately 1.056 million reads per sample. After human sequences, low quality and adapter reads were removed, a total of 7.63 billion reads were classified using a GTDB release 89 index database for taxonomic classification, resulting in 967,000 post-QC and classified reads per sample on average. In total, GTDB classification uniquely identified 151 phyla, 338 classes, 925 orders, 2,254 families, 7,906 genera and 24,705 species from gut metagenomes. We focused on common bacterial taxa to reduce alignment artefacts and noise; taxa were filtered by relative abundance (>0.01% in at least 1% of samples), which resulted in 46 phyla, 71 classes, 124 orders, 232 families, 617 genera and 1,224 species for further analysis. Overall, the most abundant taxa were members of phyla *Firmicutes, Firmicutes_A* (corresponding to *Firmicutes* in NCBI), *Firmicutes_C* (*Firmicutes*), *Bacteroidota* (*Bacteroidetes*), *Actinobacteriota* (*Actinobacteria*), and *Proteobacteria* (**Supplementary Figure 1**).

**Figure 1.**
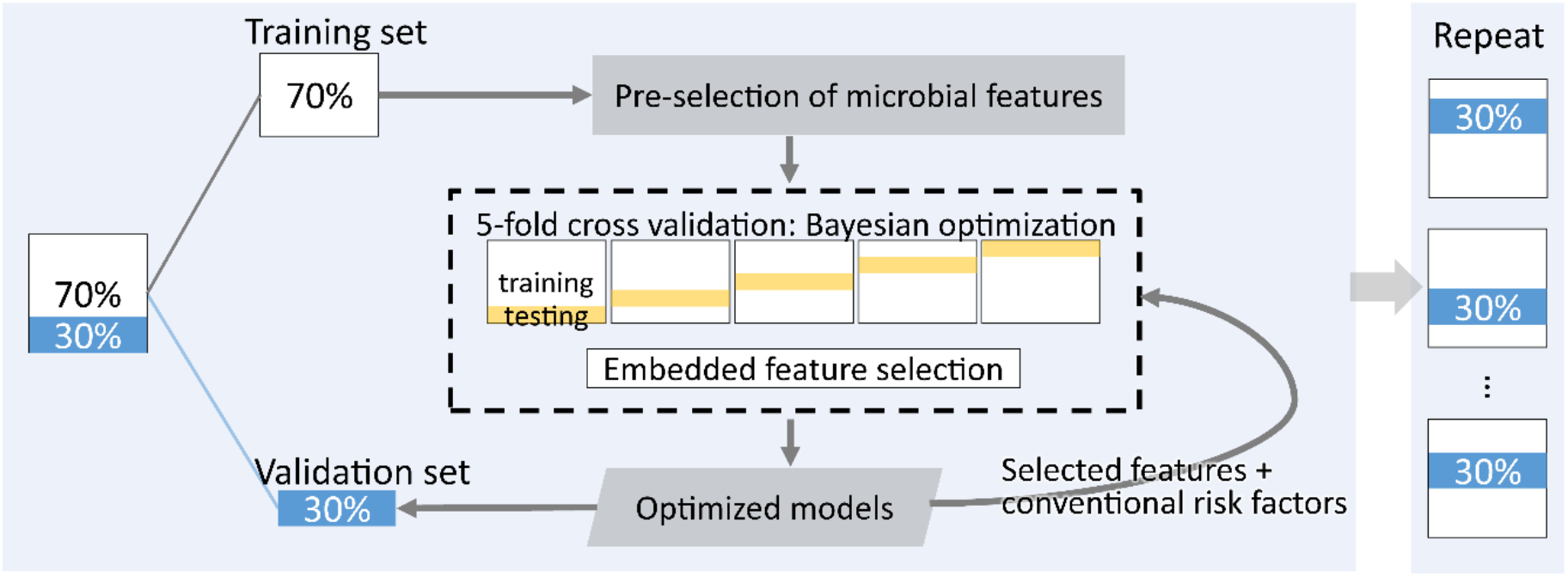
Machine learning framework for predicting incident liver disease

### Developing machine learning models

The workflow for machine learning to predict incident liver disease is shown in **Figure 1**. For both ALD and LD, samples were randomly partitioned based on the prediction target into a training set for discovery (70% of samples) and a validation set for evaluation (remaining 30%), and the partitioning itself was randomly performed 10 times to assess sampling variation. Within the training set, we developed and tested prediction models through cross-validation, and the optimal models were assessed for final performance in the withheld validation set (**Methods**). Prediction models were derived from different taxonomic levels separately, since taxa at higher ranks are inclusive of their members at lower ranks and introducing redundant features can lead to impaired prediction performance. The average results of the 10 sample partitions are reported.

To define a subset of informative taxa, we performed pre-selection of microbial features associated with incident liver disease from the union of three approaches in the training sets (**Methods**). After pre-selection, there were 10, 16, 42, 123, 355, 508 microbial taxa on average at phylum, class, order, family, genus and species levels for incident ALD, and 9, 12, 25, 62, 194, 303 for incident LD, respectively. To incorporate microbial diversity measures, Chao1, Pielou’s and Shannon’s indices were included as additional features. These microbial features were then used to build prediction models in the corresponding training sets.

Gradient boosting classifiers were applied to pre-selected microbial features to develop and optimize prediction models with cross-validation in the training datasets. To assess prediction performance, we also included two robust and common statistical approaches, logistic regression and ridge regression.

### PREDICTION OF INCIDENT LIVER DISEASE

The gradient boosting classifier generally outperformed both multivariable logistic regression and ridge regression, particularly at lower taxonomic levels (**Fig. 2**). With the gradient boosting classifier, higher prediction performance was observed at lower taxonomic levels for both incident ALD and LD, suggesting that the strength of association for higher resolution of gut microbial features outweighs their lower abundances at these levels. For LD, we obtained the highest prediction performance at species level with average AUROC of 0.733 (95% CI 0.713 - 0.752; **Fig. 2a**). At other taxonomic levels, the mean AUROC for LD ranged from 0.622 to 0.725 at phylum and genus level, respectively. When predicting ALD, we obtained average AUROC > 0.75 at phylum and class levels, and average AUROC > 0.85 for other taxonomic levels with the highest value of 0.895 (95% CI 0.881 - 0.909) at species level (**Fig. 2b**).

**Figure 2.**
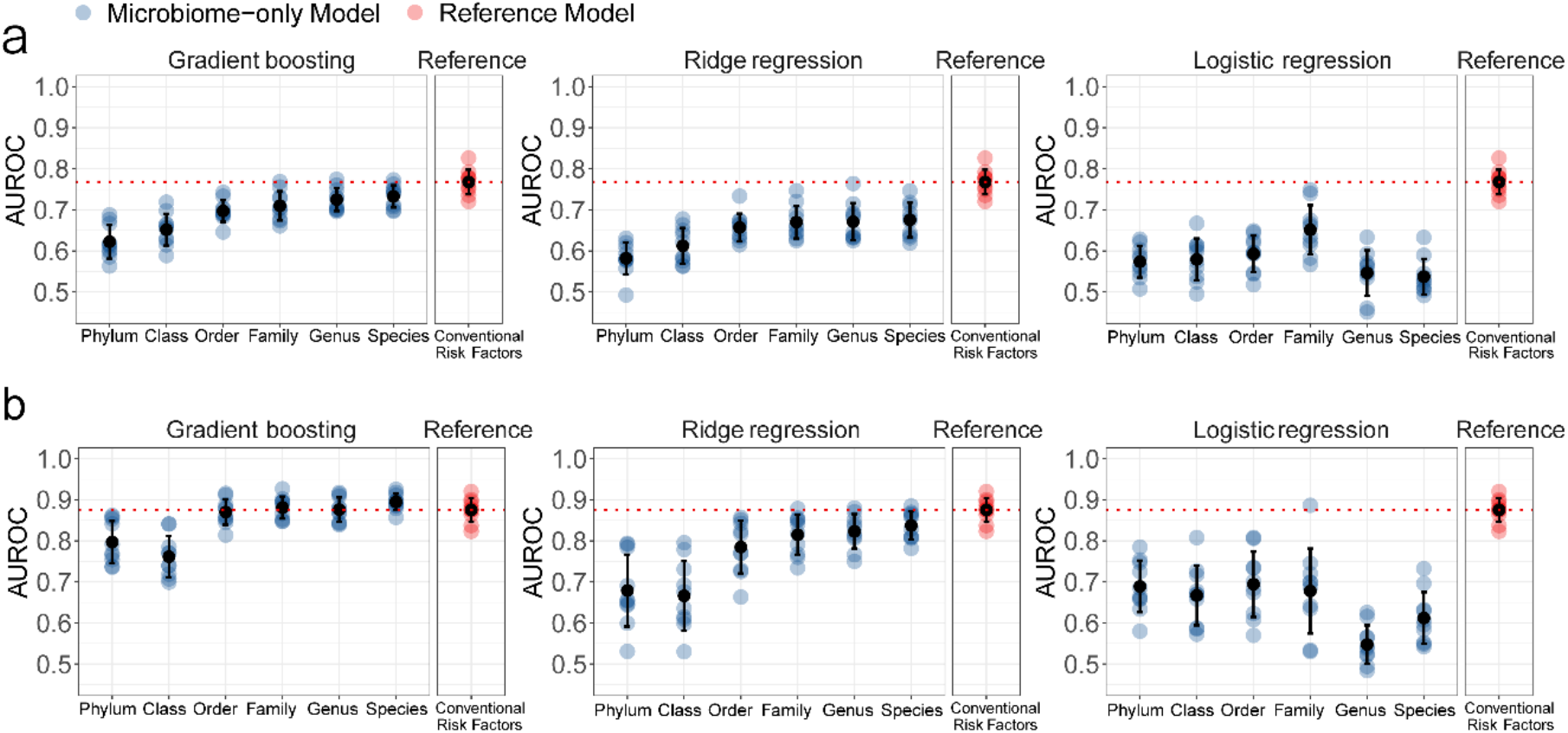
Comparison of approaches for prediction of incident liver disease using gut microbial features. **(a)** For prediction of any liver disease, the gradient boosting classifier outperformed logistic regression and ridge regression across different taxonomic levels. **(b)** For prediction of alcoholic liver disease, similar trends were observed. For comparison, a conventional prediction model is shown in red. Error bars represent mean and standard deviation. Horizontal dashed lines mark the mean performance of conventional models.

Ridge regression tended to perform better than logistic regression (**Fig. 2**). For LD, ridge regression achieved average AUROC > 0.65 at order, family, genus and species levels, with the highest AUROC of 0.675 (95% CI 0.645 - 0.706) at species level; for ALD, AUROC > 0.80 was obtained at family, genus and species levels, with the highest AUROC of 0.838 (95% CI 0.813 - 0.862) at species level. The logistic regression yielded highest AUROC of 0.651(95% CI 0.609 - 0.694) at family level and AUROC < 0.60 at other taxonomic levels for predicting LD (**Fig. 2a)**; for ALD, the best performance was obtained at order level with average AUROC of 0.694 (95% CI 0.637 - 0.751; **Fig. 2b**). Although logistic regression is highly efficient and interpretable, it did not perform well in this case where a large number of features are correlated. The L2 regularization of ridge regression better handled inter-correlated microbial features than logistic regression. However, both methods underperformed compared to the gradient boosted decision tree classifier, which is known to better capture nonlinear relationships and is robust to correlated features. The gradient boosted decision tree classifier was used in subsequent analyses.

### BENCHMARKING REFERENCE MODELS USING CONVENTIONAL APPROACHES

Conventional risk factors are commonly used for predicting liver disease risk^[20, 21]^. We built reference models using a comprehensive set of conventional risk factors, including sex, age, alcohol consumption, smoking status, body mass index (BMI), waist-hip ratio (WHR), triglycerides, high-density lipoprotein (HDL), low-density lipoprotein (LDL) and gamma-glutamyl transferase (GGT), to compare with the prediction capacity of microbiome-based models (**Methods**). The conventional prediction model achieved an average AUROC score of 0.768 (95% CI 0.746 - 0.789) for incident LD, slightly higher than the highest AUROC score of microbiome-only models achieved at species level (AUROC 0.733) (**Fig. 2a**). For ALD, the average AUROC reached 0.875 (95% CI 0.855 - 0.896), slightly lower than the AUROC of gradient boosting model achieved using species-level microbial features alone (AUROC 0.895) (**Fig. 2b**). Both conventional models and microbiome-based models had substantial predictive power individually; the next section evaluates the combination of conventional risk factors and microbial compositions for LD and ALD prediction.

### INTEGRATING GUT MICROBIOME AND CONVENTIONAL RISK FACTORS

To investigate the potential of a microbiome-augmented prediction model for liver disease, we utilised the gradient boosting classifier of microbiome features together with all conventional risk factors related to the disease, and followed the same partitioning for training and testing (**Methods**). To evaluate the performance comprehensively, the optimal models were assessed for both AUROC and AUPRC. Since greater taxonomic resolution offered better predictive performance, we compare the species-level augmented and the conventional risk factors only models.

Overall, the prediction performance of the microbiome-augmented models achieved greater AUROC and AUPRC compared with conventional prediction models. Prediction of LD (**Fig. 3a**) using the species-level augmented model yielded an average AUROC of 0.834 (95% CI 0.812 - 0.857), an AUROC increase of +0.066 over conventional prediction model (as above, average AUROC 0.768). For ALD, the species-level augmented model yielded an average AUROC of 0.956 (95% CI 0.947 - 0.965), an AUROC increase of +0.081 over conventional model (as above, average AUROC 0.875) (**Fig. 3b**).

**Figure 3.**
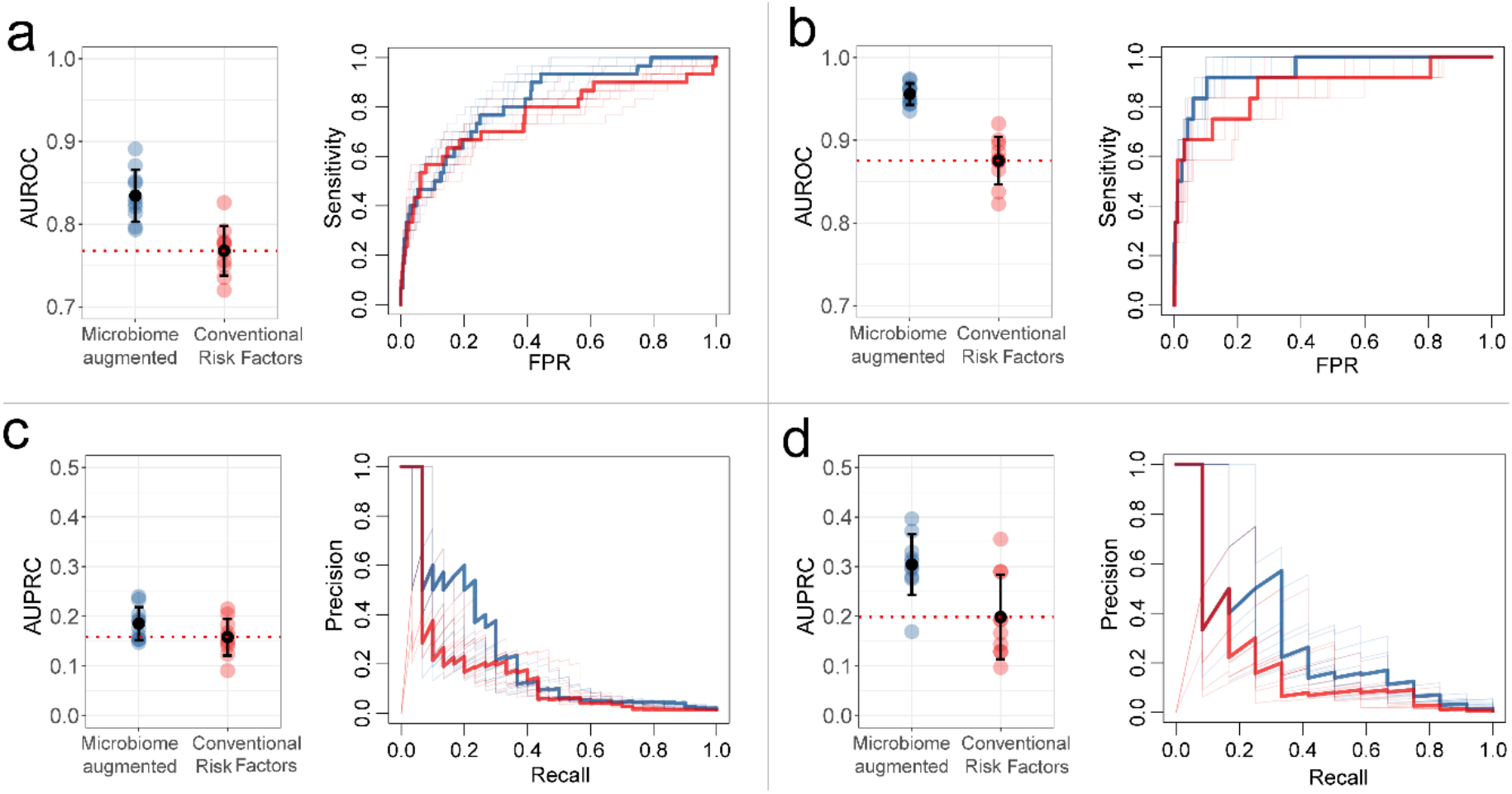
Models of conventional risk factors and gut microbiome data improved prediction of incident liver disease over conventional prediction models. Area under the ROC curve (AUROC) for gradient boosting models using species-level gut microbiome data together with conventional risk factors (blue), or a conventional risk factor model (red), with predicting **(a)** incident any liver disease or **(b)** alcoholic liver disease. Area under the precision-recall curve (AUPRC) for **(c)** any liver disease and **(d)** alcoholic liver disease. Error bars represent mean and standard deviation. Horizontal dashed lines mark the mean performance of conventional model as a reference. The bolded ROC and precision-recall curves correspond to models with AUROC and AUPRC that are closest to mean performance reference.

With a baseline AUPRC value of 0.015 for LD, the species-level augmented model achieved an average AUPRC of 0.185 (95% CI 0.161 - 0.21), which was higher than the average AUPRC of 0.158 (95% CI 0.132-0.185) yielded by the conventional prediction model (**Fig. 3c**). For ALD with a baseline AUPRC of 0.006, the species-level augmented model and conventional model achieved average AUPRC of 0.304 (95% CI 0.261 - 0.348) and 0.199 (95% CI 0.138-0.260; **Fig. 3d**), respectively.

### SURVIVAL ANALYSIS USING CONVENTIONAL AND MICROBIOME-AUGMENTED RISK MODELS

We next performed survival analysis using time-on-study Cox regression in the validation sets to assess potential clinical validity of microbiome-augmented (species level) risk models as compared to conventional risk factors only (**Methods**). The Cox model of conventional risk factors achieved average C-statistic of 0.813 (95% CI 0.792-0.835) for LD and 0.922 (95% CI 0.903-0.940) for ALD, respectively. The microbiome-augmented risk models yielded higher average c-statistic of 0.838 (95% CI 0.814-0.862) for LD and 0.959 (95% CI 0.950 - 0.968) for ALD. Consistent with this finding, the microbiome-augmented model fits significantly better (LRT p<0.01) than that using conventional risk factors only. Disease-free survival of those in the highest 5% of microbiome-augmented risk was worse than those for conventional risk factors alone (**Figure 4**).

**Figure 4.**
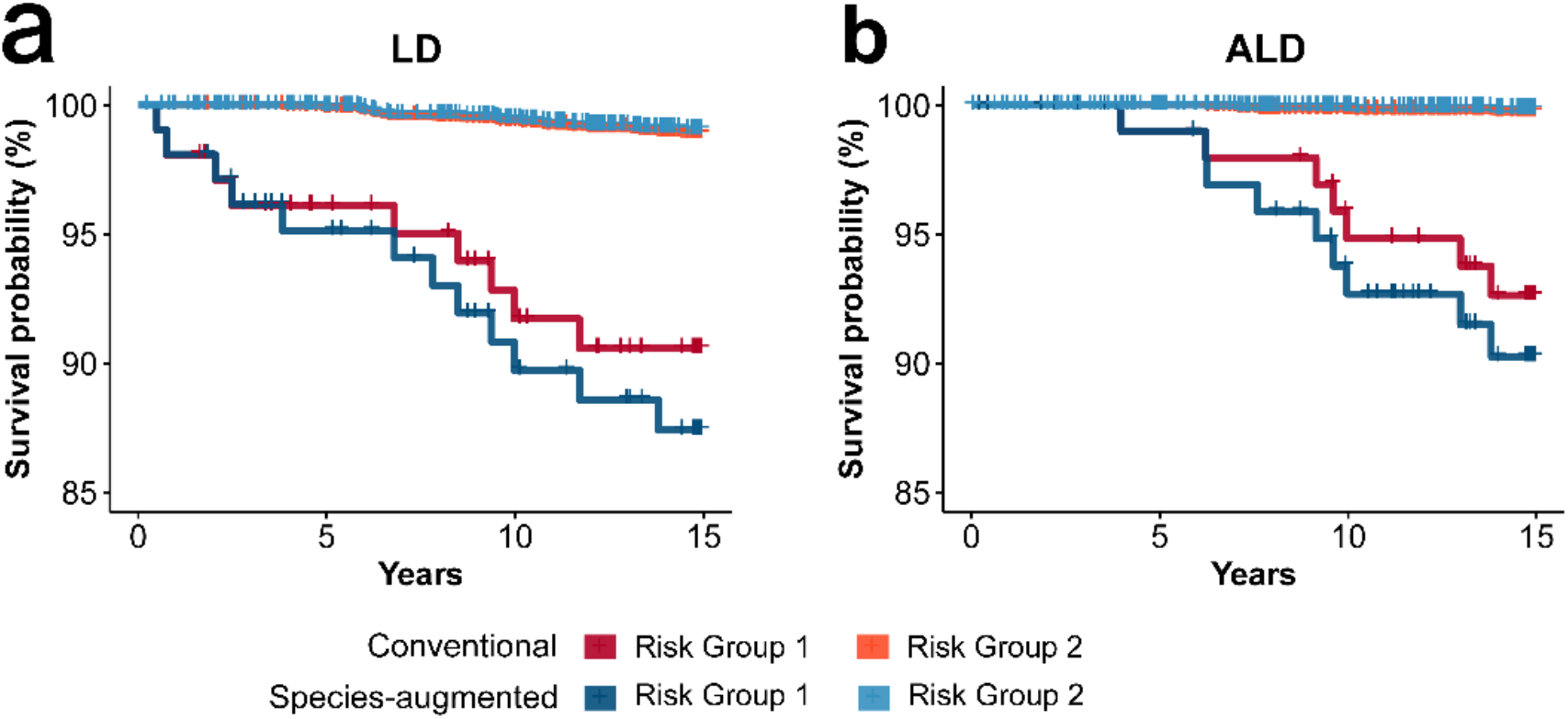
Survival curves of predicted risk groups for incident liver disease. Performance in the withheld validation set of Cox models of conventional risk factors and in combination with species-level microbiome-only scores for (a) liver disease and (b) alcoholic liver disease. Predicted risk groups are the top 5% (Risk Group 1) vs. the bottom 95% (Risk Group 2).

### COMPOSITION OF GUT MICROBIOME SIGNATURES

To better understand which bacterial taxa contribute to ALD and LD prediction, we considered those that contributed to the optimal gradient boosting classifiers at each taxonomic level, in terms of their frequency of selection and importance rank (**Supplementary Table 1**).

Notably, Pielou’s evenness, Chao1 and Shannon’s diversity, which were found to be negatively associated with both ALD and LD, were all selected as predictive contributors at phylum, class, order and family levels. This was consistent with previous findings that the richness and diversity of gut microbiome communities are positively correlated with human health^[22, 23]^.

The microbial signatures mainly comprised taxa from phylum *Actinobacteriota* (*Actinobacteria* in NCBI taxonomy), *Bacteroidota* (*Bacteroidetes*), *Firmicutes* and *Firmicutes_A* (*Firmicutes*), and *Proteobacteria* (*Proteobacteria*; **Fig. 5; Supplementary Figure 2**). Overall, most of the selected microbial taxa were significantly (FDR<0.05) and positively associated with liver disease. Many bacterial taxa have been previously reported to be related to liver disease and its progression. The families *Chitinophagaceae* (mainly contributed by *Chitinophaga*)^[24]^, *Streptococcaceae* (mainly *Streptococcus spp*.)^[24-26]^, *Enterobacteriaceae* (mainly *Klebsiella* and *Klebsiella_A*)^[25]^, genera *Actinomyces* (mainly *A. graevenitzii*)^[27, 28]^, *Rikenella*^*[29]*^, *Blautia*^*[^25, 30^]*^, *Dorea*^*[^30, 31^]*^, *Neisseria*^*[^27, 32^]*^ etc., have been frequently reported to be enriched in patients with alcoholism and ALD; the families *Streptococcaceae* (mainly *Streptococcus spp*.), *Erysipelotrichaceae, Enterobacteriaceae* (mainly *Escherichia*), genera *Actinomyces*^*[18]*^, *Lactobacillus_C* and *Lactobacillus_H* as former *Lactobacillus*^*[33-35]*^, *Veillonella*^*[^32, 34^]*^, *Prevotella spp*.^*[^13, 32, 35^-37]*^ etc., have been found to be positively associated with a broad range of liver diseases, including acute-on-chronic liver failure, non-alcoholic fatty liver disease and cirrhosis. Several members of *Actinomyces spp*.^[38, 39]^, *Escherichia spp*.^[40-42]^, *Klebsiella spp*.^[43, 44]^, *Desulfovibrio spp*.^[45]^, etc. have been identified as pathogens for liver abscess and sepsis. Consistent with previous studies, *UBA11524* (former *Faecalibacterium)*^*[^13, 18, 25, 30, 46^]*^, *Coprococcus*^*[25]*^ and *Akkermansia*^*[^25, 47^-49]*^ were negatively associated (FDR<0.05) with liver disease. Notably, genus *Akkermansia*, of which *A. muciniphila* was previously suggested as having potential protective effect on liver function and gut microbiota ecology^[48-51]^, uniquely contributed to every higher rank within phylum *Verrucomicrobia* for prediction.

**Figure 5.**
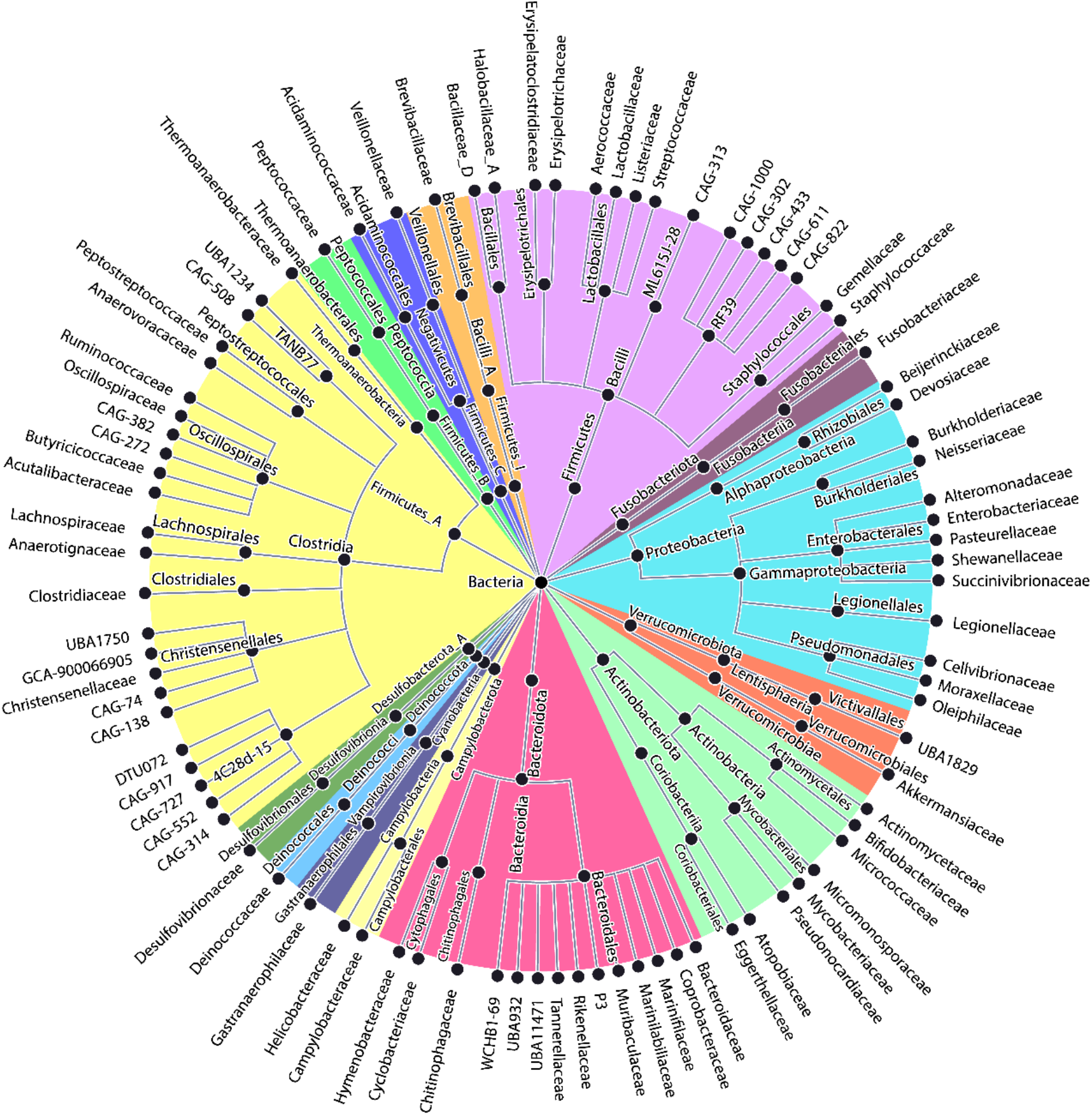
Microbial taxa predictive of liver disease. A bacterial taxonomy tree (phylum to family-level) whose members at lower ranks showed predictive signal for incident liver disease. For full taxonomy, see Supplementary Figure 2.

Among the prediction signatures, many bacterial taxa have been found in association with development of liver damage. Intestinal barrier dysfunction, marked by increased intestinal permeability, plays a key role in the pathogenesis of liver disease and is directly associated with cirrhosis^[52]^. At genus level, *Ruminococcus, Dorea, Faecalibacterium* and *Blautia* were found to be responsible for increased intestinal permeability^[53]^, which can induce translocation of microbes and microbial metabolites and subsequently worsen hepatic inflammation^[52]^. Conversely, *Bifidobacterium* was found to be negatively correlated with intestinal permeability^[53]^. Gut microbial lipopolysaccharide (LPS) is one of the most potent LPSs that triggers a cascade of proinflammatory response and promotes the progression of fatty liver^[52]^. Besides, LPS-producing bacteria are linked to obesity^[54]^, a major risk factor for NAFLD^[3]^. Although members of phylum *Bacteroidota* (*Bacteroidetes*) are largest group of LPS producers, such as *Bacteroides* and *Prevotella spp*., family *Enterobacteriaceae* of phylum *Proteobacteria* and family

*Desulfovibrionaceae* of phylum *Desulfobacterota_A* (*Proteobacteria*) exhibit an immense amount of endotoxin activity^[54]^. A recent study has shown that endotoxin-producers that overgrow in patients with fatty liver, including strain members of *Escherichia* and *Klebsiella*, can induce NAFLD in mice models and suggest a potential causative role in NAFLD^[55]^. The altered gut microbiota composition in cirrhosis is partially attributed to reduced primary bile acids and increased secondary bile acids in the gut lumen that are resulted from liver insufficiency^[52]^. The reduction of total bile acids in the gut contributes to an overgrowth of pathobiont microbes, including members of *Enterobacteriaceae* and *Enterobacteriaceae*^[52]^. The elevation of secondary bile acids is largely associated with an abundance of bacterial producers of secondary bile acid, such as members of *Clostridium* and *Eubacterium*^[52, 56]^. Bile salt hydrolase activity is associated with resistance of hepatocytes to bile toxicity and is broadly present in gut microbes including *Bacteroides, Bifidobacterium, Clostridium* and *Lactobacillus*^[56]^.

## DISCUSSION

In this study, we investigated the potential analytic and clinical validity of the gut microbiome to improve prediction of future liver disease. From baseline gut metagenomic sequencing and 15-years of EHR follow up, we developed a framework to predict incident LD and ALD using machine learning approaches, demonstrating that the gut microbiome and conventional risk factor models exhibited similar prediction performances separately, but importantly that microbiome-augmented conventional risk factor models markedly improved prediction. These results indicate that the combination of conventional risk factors with gut microbiota may have potential clinical utility in early risk stratification for liver disease.

Few studies so far have investigated the prediction of incident liver disease events using gut microbiota. Currently, clinical risk prediction models for liver disease events are commonly derived from demographic, lifestyle and biochemical factors resulted from routine blood tests. While these prediction rules have reasonable accuracy in clinical practice, they tend to be influenced by extrahepatic conditions and have reduced accuracy for early stage disease^[57, 58]^. Furthermore, there is a lack of guidance for primary care and necessity of referral based on the test results, as a large number of patients with abnormal test results are asymptomatic during liver disease progression ^[59-61]^. Thus, there is an urgent need for new tools which improve early detection of high risk individuals.

Our findings are consistent with previous studies of the relationship of bacterial taxa with hepatic function, disease and progression, and identified several with potential probiotic effects. However, the precise role of gut microbiota is poorly understood and our results support the need for species level or indeed greater levels of resolution offered by even deeper metagenomic sequencing. For example, the abundance of the *Bifidobacterium* genus has been reported to be associated with alcoholism and liver injury in various ways^[30, 62]^: at species level, *B. dentium* has been found to be enriched in advanced liver disease^[25]^, conversely *B. pseudocatenulatum* and *B. bifidum* have been recognized as potential probiotics that may attenuate liver damage^[33, 63, 64]^. This indicates the importance of lower-level taxa resolution in interpreting how bacteria contribute to the disease pathology.

Our study has several limitations. Due to the necessity of a prospective early detection study to consider a large number of apparently healthy individuals, we were limited in the number of incident disease cases, and therefore we are not well-powered to investigate subtypes and stages of liver disease which might lead to greater clinical significance. The need for shallow metagenomic sequencing for a large prospective cohort also meant that we were not able to evaluate the added information of deep sequencing to risk prediction. The prevention measures available to individuals at high risk of liver disease are also somewhat limited. These include weight reduction, alcohol and smoking cessation, and may extend to caution with pharmaceutical prescriptions. Finally, our cohort is of European ancestry and therefore likely suffers from the well-known ancestry bias of analyses performed in European cohorts; thus, these prediction models are likely to have attenuated performance in non-European ancestries.

Notwithstanding the challenging necessity for validation of novel biomarkers as well as development of standards for interpretation as prerequisites for clinical implementation, our study provides an evidence base and corresponding risk prediction models for the translation of metagenomic sequencing in risk prediction of liver disease.

## METHODS

### STUDY POPULATION

The FINRISK population surveys have been performed every 5 years since 1972 to monitor trends in cardiovascular disease risk factors in the Finnish population^[19, 65]^. The FINRISK 2002 study was based on a stratified random sample of the population aged 25–74 years from six specific geographical areas of Finland^[66]^. The sampling was stratified by sex, region and 10-year age group so that each stratum had 250 participants. The overall participation rate was 65.5% (n = 8798). The participants filled out a questionnaire at home, then participated in a clinical examination carried out by specifically trained nurses and gave a blood sample. They also received a sampling kit and instructions to donate a stool sample at home and mail it to the Finnish Institute for Health and Welfare in an overnight mail. The follow up of the cohort took place by record linkage of the study data with the Finnish national electronic health registers (Hospital Discharge Register and Causes of Death Register), which provide in practice 100% coverage of relevant health events in Finnish residents. For the present analyses the follow-up extended until Dec 31^st^, 2016. The study protocol of FINRISK 2002 was approved by the Coordinating Ethical Committee of the Helsinki and Uusimaa Hospital District (Ref. 558/E3/2001). All participants signed an informed consent. The study was conducted according to the World Medical Association’s Declaration of Helsinki on ethical principles.

### PHENOTYPE METADATA

The phenotype data in this study comprised of demographic characteristics, life habits, disease history and medications, laboratory test results and follow-up EHRs. Baseline phenotype variables used as conventional risk factors included age, sex, body mass index (BMI), waist-hip ratio (WHR), smoking status, alcoholic consumption, triglyceride (TRIG), gamma-glutamyl transferase (GGT), high-density lipoprotein (HDL) and low-density lipoprotein (LDL) cholesterol measurements. BMI was computed as the weight in kilograms divided by the square of height in meters measure with light clothing^[19]^. Smoking status described whether a participant was a current daily smoker at the time of the survey. Alcohol consumption, based on self-reported questionnaire, was measured as the average weekly pure alcohol use in grams during the past 12 months. TRIG, GGT, HDL and LDL-cholesterol were measured from blood samples collected from participants advised to fast for at least 4 hours prior to collection and avoid heavy meals earlier during the day^[19, 67, 68]^. EHR follow-up of incident disease was until December 31^st^, 2016. The median follow-up was 14.84 years and the end point was the date of death or last follow-up. Incident disease was coded as a binary variable indicating disease case (1) or non-case (0) with matched time from baseline to event or end of follow-up also utilised for analyses.

### CHARACTERIZATION OF THE GUT MICROBIOME

Stool samples were collected by participants and mailed overnight to Finnish Institute for Health and Welfare for storing at −20°C; the samples were sequenced at the University of California San Diego in 2017. The gut microbiome was characterized by shallow shotgun metagenomics sequencing with Illumina HiSeq 4000 Systems. We successfully performed stool shotgun sequencing in n = 7231 individuals. The detailed procedures for DNA extraction, library preparation and sequence processing have been previously described^[66]^. Adapter and host sequences were removed. To preserve the quality of data while retaining most of the disease cases, samples with sequencing depth less than 400,000 were excluded from our analysis. The metagenomes were classified using default parameters in Centrifuge 1.0.4^[69]^, and using an index database based on taxonomic definitions from the Genome Taxonomy Database (GTDB) release 89^[70] [71]^.

The gut microbial composition was represented as relative abundance of taxa. For each metagenome at phylum, class, order, family, genus and species levels, the relative abundance of a taxon was computed as the proportion of reads assigned to the clade rooted at this taxon among total classified reads of this metagenome. The relative abundance of a bacteria that had no reads assigned in a metagenome was considered as zero in the corresponding profile. We focused on common and relatively abundant taxa of a within-sample relative abundance greater than 0.01% in more than 1% of samples. The centered log-ratio (CLR) transformation was carried out on abundance data by taking the log of taxa abundance divided by geometric mean of abundance in each metagenome profile. Abundance of zero was replaced with a value representing 1/10 of the minimum abundance in a metagenome before transformation. In this study, all analyses except for microbial diversity calculation were based on CLR transformed data.

### DISEASE CASE DEFINITIONS

The liver disease investigated in this study consists of two groups, alcoholic liver disease (ALD) and a broader range of any liver disease (LD) according to the ICD-10 codes (Finnish modification). A sample was considered as an incident case of any liver disease if the follow-up register-based diagnostic classification was under the ICD-10 codes K70 - K77; the alcoholic liver disease was defined by the ICD-10 code K70. In the present study, the disease diagnosis was last followed up by the end of 2016.

### INCLUSION AND EXCLUSION CRITERIA

The inclusion criteria of FINRISK 2002 cohort have been previously described^[19]^. Samples with gut microbiome profiles, phenotype metadata and follow-up all available were included in our analysis (n=7115). The exclusion criteria of our analysis were: (1) samples with gut metagenomic sequencing yielding <400K reads; (2) presence of baseline prevalent diagnosis of target disease for prediction; (3) baseline pregnancy during the survey year. Altogether, 7005 and 6965 samples were included for modelling analyses of ALD and LD, respectively.

### PREDICTION MODELING OF INCIDENT LIVER DISEASES

#### General framework

Prediction models were developed for any liver disease and alcoholic liver disease at phylum, class, order, family, genus and species levels separately. For each incident disease to be predicted, samples were randomly shuffled and partitioned into a training cohort for discovery and a validation cohort for evaluation at a 7:3 ratio according to the target disease variable such that the distribution of disease cases and healthy controls in training and testing datasets were consistent. Within the training set, we first performed pre-selection of features (detailed in next section) and then developed models using pre-selected features through 5-fold cross validation stratified according to the prediction target, which further created random splits of internal training and testing sets at a 8:2 ratios five times with testing sets being mutually exclusive. The models were optimized based on cross-validated results. The optimal models were then trained on the full training set and finally assessed on the withheld validation set that was excluded from the training and optimization process to avoid data leakage from the training set. Considering the variation of attribute distributions that can occur during random data partitioning, we repeated the whole process described above 10 times and reported the average results. The detailed procedures were elucidated in the rest of this section.

#### Pre-selection of microbial taxa

To select a set of informative microbial taxa that were individually associated with incident liver disease, we analyzed the relationship between microbial abundance and incident disease using (1) logistic regression adjusted for age and gender, (2) Cox regression for time to disease occurrence adjusted for age and gender, and (3) Spearman correlation. This feature selection step was performed only within the training datasets accounting for 70% of samples. A microbial taxon was included in further analyses if statistical significance (P<0.05) was found by any of the above three methods. After adjusting for age and gender, on average 8 phyla, 14 classes, 35 orders, 103 families, 299 genus and 406 species were associated with incident ALD at statistical significance using logistic regression; 8 phyla, 14 classes, 36 orders, 106 families, 306 genera and 416 species were found significant using cox regression. The Spearman correlation found 7 phyla, 12 classes, 36 orders, 112 families, 314 genera and 428 species, on average, significantly correlated with alcoholic liver disease. For LD, the average numbers of significantly associated taxa at each taxonomic level were 7 phyla, 10 classes, 19 orders, 49 families, 157 genera and 245 species with logistic regression; 7 phyla, 10 classes, 20 orders, 52 families, 164 genera and 255 species with cox regression; 5 phyla, 8 classes, 19 orders, 51 families, 148 genera and 218 species with Spearman correlation. As the selected taxa were not always agreed by all three approaches, taxa selected by any approach in the training cohort were included for developing prediction models with the corresponding data partition. Of the 10 differently sampled training sets, the average numbers of microbial features at phylum, class, order, family, genus and species levels were 10, 16, 42, 123, 355, 508 for predicting incident ALD, and 9, 12, 25, 62, 194, 303 for predicting LD, respectively.

#### Microbial and conventional features

Conventional risk factors include baseline age, gender, BMI (kg/m^2^), WHR, alcohol consumption (g), smoking status, TRIG (mmol/l), GGT (U/L), HDL and LDL cholesterol (mmol/l). Microbial features comprised taxa abundance along with microbial diversity metrics at phylum, class, order, family, genus and species levels. To characterize microbial diversity in samples, Chao1 index, Pielou’s evenness index and Shannon diversity index were calculated using raw abundance data without filtering. Chao1 index estimates the total species richness for a given community considering the presence of rare species. Pielou’s evenness index measures how evenly the species are distributed in a given sample. Shannon’s index takes into account both species richness and evenness.

#### Model development

The machine learning approach extreme gradient boosting was applied to predict the incidence of liver disease from baseline phenotype and microbial data using *Xgboost* library in R. Xgboost is a distributed and optimized implementation of gradient boosting decision trees, an ensemble method of sequential and additive training of trees with regularizations^[72]^. The prediction procedure was a twofold process which involved developing models using microbial features alone and in combination with conventional risk factors. In the first step the gradient boosting classifiers were trained on microbial features consisting of taxa abundance and diversity metrics at different taxonomic levels separately. In the second step, microbial features selected by the embedded feature selection of gradient boosting classifiers in the first step, together with conventional risk factors, were deployed to predict incident disease. The models were trained with Bayesian optimization (*mlrMBO* in R) through 5-fold cross validation in the training dataset. The optimal models selected based on cross-validated results were evaluated in the withheld evaluation dataset as the final performance for predicting incident disease. The highly ranked and frequently selected (by more than half of the models) microbial features were considered as predictive signatures for further interpretation. Since logistic regression was one of the most widely used statistical tools for building clinical prediction models, we compared its prediction performance with gradient boosting classifiers using the same training and evaluation sets. In addition, we performed Ridge regression, which was more suited to correlated microbiome features by adding an L2 penalty term to the loss function, following consistent data partitioning strategies. The Ridge regression was optimized by a fine grid search of parameters with cross-validation of the same divisions of folds as the gradient boosting classifier.

#### Benchmarking reference models with conventional methods

Currently, prediction models for liver disease are commonly built by regression methods of conventional risk factors. Therefore, reference models were built using logistic regression of commonly used liver disease predictors including age, gender, BMI (kg/m^2^), WHR, alcoholic consumption (g), smoking status, TRIG (mmol/l), GGT (U/L), HDL and LDL cholesterol (mmol/l), as a benchmark procedure.

#### Model evaluation

The prediction performance of all models was evaluated in the corresponding withheld validation dataset (30% of samples) that were not used for discovery. The area under the receiver operating characteristic curve (AUROC) was used to compare the performance across models of different methods and features. The AUROC is a widely applied metric that considers the trade-offs between sensitivity and specificity at all possible thresholds for comparing the performance across various classifiers with a baseline value of 0.5 for a random classifier. Area under the precision-recall curve (AUPRC) was provided as a complementary assessment, particularly when constructing risk models combining microbiome and conventional risk factors. AUPRC considers the trade-offs between precision (or positive predictive value) and recall (or sensitivity) with a baseline that equals the proportion of positive disease cases in all samples. Since AUPRC is more sensitive to higher ranks of the positive class, it is preferred for highly imbalanced datasets where, for example, case numbers are small relative to controls. As the entire model development process was repeated 10 times, following the 10 randomly sampled partitions of training and validation datasets, each data partitioning led to a set of optimal models developed in the corresponding training dataset. The final performance of optimal models developed from discovery data was evaluated in the corresponding validation data that were set apart in the beginning. The average results of data partitions were reported. To further assess the final prediction result, we considered the species-level microbiome models using gradient boosting classifiers, which outperformed microbiome-only models based on other taxonomic levels for both LD and ALD. In the withheld validation datasets of various partitions, Cox regression models of conventional predictors and in combination with predicted scores of microbiome-only models were built using the time difference between baseline and follow-up disease occurrence or the end of follow-up. The Cox models were evaluated by the concordance statistic (c-statistic). The fit of the model was assessed by likelihood ratio test.

### Data Availability

The data for the present study are available with a written application to the THL Biobank as instructed in the website of the Biobank: https://thl.fi/en/web/thl-biobank/for-researchers. Predictive models are available at https://doi.org/10.26188/12554573.v1.

## ACKNOWLEDGEMENTS

VS was supported by the Finnish Foundation for Cardiovascular Research. MI was supported by the Munz Chair of Cardiovascular Prediction and Prevention. ASH was supported by the Academy of Finland, grant no. 321356. LL was supported by Academy of Finland (295741, 307127). TN was supported by the Emil Aaltonen Foundation, the Paavo Nurmi Foundation, the Finnish Medical Foundation, and the Academy of Finland (grant no. 321351). RL receives funding support from NIEHS (5P42ES010337), NCATS (5UL1TR001442), NIDDK (U01DK061734, R01DK106419, P30DK120515, R01DK121378, R01DK124318), and DOD PRCRP (W81XWH-18-2-0026). This study was supported by the Victorian Government’s Operational Infrastructure Support (OIS) program, and by core funding from: the UK Medical Research Council (MR/L003120/1), the British Heart Foundation (RG/13/13/30194; RG/18/13/33946) and the National Institute for Health Research [Cambridge Biomedical Research Centre at the Cambridge University Hospitals NHS Foundation Trust] [*]. This work was supported by Health Data Research UK, which is funded by the UK Medical Research Council, Engineering and Physical Sciences Research Council, Economic and Social Research Council, Department of Health and Social Care (England), Chief Scientist Office of the Scottish Government Health and Social Care Directorates, Health and Social Care Research and Development Division (Welsh Government), Public Health Agency (Northern Ireland), British Heart Foundation and Wellcome.

*The views expressed are those of the authors and not necessarily those of the NHS, the NIHR or the Department of Health and Social Care.

## References

1. World health statistics overview 2019: monitoring health for the SDGs, sustainable development goals.. Licence: CC BY-NC-SA 3.0 IGO ed. Vol. (WHO/DAD/2019.1). 2019, Geneva: World Health Organization.

2. Asrani, S.K., et al., Burden of liver diseases in the world. J Hepatol, 2019. 70(1): p. 151–171.

3. Younossi, Z., et al., Global burden of NAFLD and NASH: trends, predictions, risk factors and prevention. Nat Rev Gastroenterol Hepatol, 2018. 15(1): p. 11–20.

4. Younossi, Z.M., et al., Global epidemiology of nonalcoholic fatty liver disease-Meta-analytic assessment of prevalence, incidence, and outcomes. Hepatology, 2016. 64(1): p. 73–84.

5. Bellentani, S., The epidemiology of non-alcoholic fatty liver disease. Liver Int, 2017. 37 Suppl 1: p. 81–84.

6. Soresi, M., et al., Non invasive tools for the diagnosis of liver cirrhosis. World J Gastroenterol, 2014. 20(48): p. 18131–50.

7. Cleveland, E., A. Bandy, and L.B. VanWagner, Diagnostic challenges of nonalcoholic fatty liver disease/nonalcoholic steatohepatitis. Clin Liver Dis (Hoboken), 2018. 11(4): p. 98–104.

8. Moreno, C., S. Mueller, and G. Szabo, Non-invasive diagnosis and biomarkers in alcohol-related liver disease. J Hepatol, 2019. 70(2): p. 273–283.

9. Hartmann, P., et al., Gut microbiota in liver disease: too much is harmful, nothing at all is not helpful either. Am J Physiol Gastrointest Liver Physiol, 2019. 316(5): p. G563–G573.

10. Tripathi, A., et al., The gut-liver axis and the intersection with the microbiome. Nat Rev Gastroenterol Hepatol, 2018. 15(7): p. 397–411.

11. Adolph, T.E., et al., Liver-Microbiome Axis in Health and Disease. Trends Immunol, 2018. 39(9): p. 712–723.

12. Safari, Z. and P. Gérard, The links between the gut microbiome and non-alcoholic fatty liver disease (NAFLD). Cellular and Molecular Life Sciences, 2019. 76(8): p. 1541–1558.

13. Zhu, L., R.D. Baker, and S.S. Baker, Gut microbiome and nonalcoholic fatty liver diseases. Pediatr Res, 2015. 77(1-2): p. 245–51.

14. Szabo, G., Gut-liver axis in alcoholic liver disease. Gastroenterology, 2015. 148(1): p. 30–6.

15. Woodhouse, C.A., et al., Review article: the gut microbiome as a therapeutic target in the pathogenesis and treatment of chronic liver disease. Alimentary Pharmacology & Therapeutics, 2018. 47(2): p. 192–202.

16. Qin, N., et al., Alterations of the human gut microbiome in liver cirrhosis. Nature, 2014. 513(7516): p. 59–64.

17. Loomba, R., et al., Gut Microbiome-Based Metagenomic Signature for Non-invasive Detection of Advanced Fibrosis in Human Nonalcoholic Fatty Liver Disease. Cell Metab, 2017. 25(5): p. 1054–1062 e5.

18. Caussy, C., et al., A gut microbiome signature for cirrhosis due to nonalcoholic fatty liver disease. Nature Communications, 2019. 10(1): p. 1406.

19. Borodulin, K., et al., Cohort Profile: The National FINRISK Study. Int J Epidemiol, 2018. 47(3): p. 696–696i.

20. Bedogni, G., et al., The Fatty Liver Index: a simple and accurate predictor of hepatic steatosis in the general population. BMC gastroenterology, 2006. 6: p. 33–33.

21. Long, M.T., et al., A simple clinical model predicts incident hepatic steatosis in a community-based cohort: The Framingham Heart Study. Liver international : official journal of the International Association for the Study of the Liver, 2018. 38(8): p. 1495–1503.

22. Acharya, C. and J.S. Bajaj, Gut Microbiota and Complications of Liver Disease. Gastroenterology clinics of North America, 2017. 46(1): p. 155–169.

23. Backhed, F., et al., Defining a healthy human gut microbiome: current concepts, future directions, and clinical applications. Cell Host Microbe, 2012. 12(5): p. 611–22.

24. Puri, P., et al., The circulating microbiome signature and inferred functional metagenomics in alcoholic hepatitis. Hepatology, 2018. 67(4): p. 1284–1302.

25. Dubinkina, V.B., et al., Links of gut microbiota composition with alcohol dependence syndrome and alcoholic liver disease. Microbiome, 2017. 5(1): p. 141.

26. Sarin, S.K., A. Pande, and B. Schnabl, Microbiome as a therapeutic target in alcohol-related liver disease. Journal of Hepatology, 2019. 70(2): p. 260–272.

27. Fan, X., et al., Drinking alcohol is associated with variation in the human oral microbiome in a large study of American adults. Microbiome, 2018. 6(1): p. 59–59.

28. Hwang, S.S., et al., Actinomyces graevenitzii bacteremia in a patient with alcoholic liver cirrhosis. Anaerobe, 2011. 17(2): p. 87–89.

29. Xue, M., et al., Protective effect of aplysin on liver tissue and the gut microbiota in alcohol-fed rats. PloS one, 2017. 12(6): p. e0178684–e0178684.

30. Leclercq, S., et al., Intestinal permeability, gut-bacterial dysbiosis, and behavioral markers of alcohol-dependence severity. Proceedings of the National Academy of Sciences of the United States of America, 2014. 111(42): p. E4485–E4493.

31. Waters, J.L. and R.E. Ley, The human gut bacteria Christensenellaceae are widespread, heritable, and associated with health. BMC Biology, 2019. 17(1): p. 83.

32. Chen, Y., et al., Dysbiosis of small intestinal microbiota in liver cirrhosis and its association with etiology. Scientific reports, 2016. 6: p. 34055-34055.

33. Nobili, V., et al., Bifidobacteria and lactobacilli in the gut microbiome of children with non-alcoholic fatty liver disease: which strains act as health players? Archives of medical science : AMS, 2018. 14(1): p. 81–87.

34. Shao, L., et al., Disorganized Gut Microbiome Contributed to Liver Cirrhosis Progression: A Meta-Omics-Based Study. Frontiers in microbiology, 2018. 9: p. 3166–3166.

35. Jiang, W., et al., Dysbiosis gut microbiota associated with inflammation and impaired mucosal immune function in intestine of humans with non-alcoholic fatty liver disease. Scientific Reports, 2015. 5(1): p. 8096.

36. Shen, F., et al., Gut microbiota dysbiosis in patients with non-alcoholic fatty liver disease. Hepatobiliary & Pancreatic Diseases International, 2017. 16(4): p. 375–381.

37. Boursier, J., et al., The severity of nonalcoholic fatty liver disease is associated with gut dysbiosis and shift in the metabolic function of the gut microbiota. Hepatology (Baltimore, Md.), 2016. 63(3): p. 764–775.

38. Ávila, F., et al., Hepatic Actinomycosis. GE Portuguese journal of gastroenterology, 2015. 22(1): p. 19–23.

39. Könönen, E. and W.G. Wade, Actinomyces and related organisms in human infections. Clinical microbiology reviews, 2015. 28(2): p. 419–442.

40. Yamamoto, D., et al., Escherichia albertii, a novel human enteropathogen, colonizes rat enterocytes and translocates to extra-intestinal sites. PloS one, 2017. 12(2): p. e0171385–e0171385.

41. Commander, S.J., et al., Liver abscesses secondary to Escherichia coli infection mimicking multifocal hepatoblastoma: A case report. Journal of Pediatric Surgery Case Reports, 2017. 18: p. 42–44.

42. Chen, S.-C., et al., Pyogenic liver abscesses with Escherichia coli: etiology, clinical course, outcome, and prognostic factors. Wiener klinische Wochenschrift, 2005. 117(23): p. 809–815.

43. Paasch, C., S. Wilczek, and M.W. Strik, Liver abscess and sepsis caused by Clostridium perfringens and Klebsiella oxytoca. International journal of surgery case reports, 2017. 41: p. 180–183.

44. Kamal, F., et al., Klebsiella Pneumoniae Liver Abscess: a Case Report and Review of Literature. Cureus, 2017. 9(1): p. e970–e970.

45. Koyano, S., et al., A Case of Liver Abscess with Desulfovibrio desulfuricans Bacteremia. Case reports in infectious diseases, 2015. 2015: p. 354168–354168.

46. Yun, Y., et al., Fecal and blood microbiota profiles and presence of nonalcoholic fatty liver disease in obese versus lean subjects. PloS one, 2019. 14(3): p. e0213692–e0213692.

47. Lowe, P.P., et al., Alcohol-related changes in the intestinal microbiome influence neutrophil infiltration, inflammation and steatosis in early alcoholic hepatitis in mice. PloS one, 2017. 12(3): p. e0174544–e0174544.

48. Grander, C., et al., Recovery of ethanol-induced *Akkermansia muciniphila* depletion ameliorates alcoholic liver disease. Gut, 2018. 67(5): p. 891.

49. Wu, W., et al., Protective Effect of Akkermansia muciniphila against Immune-Mediated Liver Injury in a Mouse Model. Frontiers in microbiology, 2017. 8: p. 1804-1804.

50. Dao, M.C., et al., *Akkermansia muciniphila* and improved metabolic health during a dietary intervention in obesity: relationship with gut microbiome richness and ecology. Gut, 2016. 65(3): p. 426.

51. Kim, S., et al., *Akkermansia muciniphila* Prevents Fatty Liver, Decreases Serum Triglycerides, and Maintains Gut Homeostasis. Applied and Environmental Microbiology, 2020: p. AEM.03004-19.

52. Albillos, A., A. de Gottardi, and M. Rescigno, The gut-liver axis in liver disease: Pathophysiological basis for therapy. J Hepatol, 2020. 72(3): p. 558–577.

53. Leclercq, S., et al., Intestinal permeability, gut-bacterial dysbiosis, and behavioral markers of alcohol-dependence severity. Proc Natl Acad Sci U S A, 2014. 111(42): p. E4485–93.

54. Zhao, L., The gut microbiota and obesity: from correlation to causality. Nat Rev Microbiol, 2013. 11(9): p. 639–47.

55. Fei, N., et al., Endotoxin Producers Overgrowing in Human Gut Microbiota as the Causative Agents for Nonalcoholic Fatty Liver Disease. mBio, 2020. 11(1): p. e03263–19.

56. Wahlström, A., et al., Intestinal Crosstalk between Bile Acids and Microbiota and Its Impact on Host Metabolism. Cell Metab, 2016. 24(1): p. 41–50.

57. Vilar-Gomez, E. and N. Chalasani, Non-invasive assessment of non-alcoholic fatty liver disease: Clinical prediction rules and blood-based biomarkers. J Hepatol, 2018. 68(2): p. 305–315.

58. Carbone, M., et al., The UK-PBC risk scores: Derivation and validation of a scoring system for long-term prediction of end-stage liver disease in primary biliary cholangitis. Hepatology, 2016. 63(3): p. 930–50.

59. Williams, R., et al., Disease burden and costs from excess alcohol consumption, obesity, and viral hepatitis: fourth report of the Lancet Standing Commission on Liver Disease in the UK. Lancet, 2018. 391(10125): p. 1097–1107.

60. Standing, H.C., et al., GPs’ experiences and perceptions of early detection of liver disease: a qualitative study in primary care. Br J Gen Pract, 2018. 68(676): p. e743–e749.

61. Harmala, S., et al., Development and validation of a prediction model to estimate the risk of liver cirrhosis in primary care patients with abnormal liver blood test results: protocol for an electronic health record study in Clinical Practice Research Datalink. Diagn Progn Res, 2019. 3: p. 10.

62. Xu, M., et al., Changes of Fecal Bifidobacterium Species in Adult Patients with Hepatitis B Virus-Induced Chronic Liver Disease. Microbial Ecology, 2012. 63(2): p. 304–313.

63. Fang, D., et al., Bifidobacterium pseudocatenulatum LI09 and Bifidobacterium catenulatum LI10 attenuate D-galactosamine-induced liver injury by modifying the gut microbiota. Scientific Reports, 2017. 7(1): p. 8770.

64. Gómez-Hurtado, I., et al., Improved hemodynamic and liver function in portal hypertensive cirrhotic rats after administration of B. pseudocatenulatum CECT 7765. European Journal of Nutrition, 2019. 58(4): p. 1647–1658.

65. Borodulin, K., et al., Forty-year trends in cardiovascular risk factors in Finland. Eur J Public Health, 2015. 25(3): p. 539–46.

66. Salosensaari, A., et al., Taxonomic Signatures of Long-Term Mortality Risk in Human Gut Microbiota. medRxiv, 2020:. 2019.12.30.19015842.

67. Juutilainen, A., et al., Trends in estimated kidney function: the FINRISK surveys. Eur J Epidemiol, 2012. 27(4): p. 305–13.

68. Havulinna, A.S., et al., Circulating Ceramides Predict Cardiovascular Outcomes in the Population-Based FINRISK 2002 Cohort. Arterioscler Thromb Vasc Biol, 2016. 36(12): p. 2424–2430.

69. Kim, D., et al., Centrifuge: rapid and sensitive classification of metagenomic sequences. Genome Res, 2016. 26(12): p. 1721–1729.

70. Parks, D.H., et al., A standardized bacterial taxonomy based on genome phylogeny substantially revises the tree of life. Nat Biotechnol, 2018. 36(10): p. 996–1004.

71. Wick, R. and G. Méric, Metagenomics Index Correction. 2019.

72. Chen, T. and C. Guestrin, XGBoost: A Scalable Tree Boosting System, in KDD ‘16: Proceedings of the 22Nd ACM SIGKDD International Conference on Knowledge Discovery and Data Mining. 2016, ACM. p. 785–794.

